# Reciprocal association between participation to a national election and the epidemic spread of COVID-19 in France: nationwide observational and dynamic modeling study

**DOI:** 10.1101/2020.05.14.20090100

**Authors:** Jean-David Zeitoun, Matthieu Faron, Sylvain Manternach, Jérôme Fourquet, Marc Lavielle, Jérémie H. Lefèvre

## Abstract

**Objective:** To investigate possible reciprocal associations between the intensity of the COVID-19 epidemic in France and the level of participation at national elections.

**Design:** Observational study and dynamic modelling using a sigmoidal mixed effects model.

**Setting:** All hospitals where patients were admitted for COVID-19.

**Participants:** All admitted patients from March 18, 2020 to April 17, 2020.

**Main outcome measures:** Abstention and admission rate for COVID-19.

**Results:** Mean abstention rate in 2020 among departments was 52.5%±6.4 and had increased by a mean of 18.8% as compared with the 2014 election. There was a high degree of similarity of abstention between the two elections among the departments (p<0.001). Among departments with a high outbreak intensity before the election, those with a higher participation were not affected by a significantly higher number of COVID-19 admissions after the elections. The sigmoidal model fitted the data from the different departments with a high degree of consistency. The covariate analysis showed that a significant association between participation and number of admitted patients was observed for both elections (2020: β=-5.36, p<1e^−9^ and 2014: β=-3.15, p<1e^−6^) contradicting a direct specific causation of the 2020 election. Participation was not associated with the position of the inflexion point suggesting no effect in the speed of spread.

**Conclusions:** Our results suggest that the surrounding intensity of the COVID-19 epidemic in France did not have any local impact on citizens’ participation to a national election. The level of participation to the 2020 election had no impact on the spread of the pandemic.

## Introduction

The outbreak of coronavirus disease 19 (COVID-19), the viral pneumonia related to severe acute respiratory syndrome coronavirus 2 (SARS-CoV-2), has led the French government to impose a mandatory national containment. [1] It was officially advertised by the French President on March 12, 2020 yet containment was scheduled to be fully effective as of March 17, 2020. The first round of the municipal elections for mayor designation in all French cities was to be held on March 15, 2020, i.e. between the announcement and the implementation of the containment, while the phase III of the epidemic has been declared on March 14, 2020. The governmental decision to maintain those elections has been a matter of national controversy. [2] Containment had already been decided and defined as a protective public health measure, and many stakeholders claimed that barrier gestures and other countermeasures would not be properly applied along the event in every voting office. The elections took place, yet national participation was historically low as compared to prior occurrences of the same types of elections. Anecdotal reports suggested that some city officials were affected by COVID-19 or even died from it within days or weeks following the elections. [3] However, whether the people participation was influenced by the surrounding intensity of epidemic has not been analyzed. On the other hand, whether the holding of those elections has actually been a worsening factor increasing the spread of the epidemic is unknown.

We sought to answer these two questions by examining the pattern of COVID-19 spread before and in the wake of the elections, and by studying its possible association with the level of voting participation.

## Methods

We retrieved demographic characteristics regarding the 95 French departments, which were our geographic unit of analysis: number of inhabitants and surface. We also collected data regarding the epidemic around the time of the elections and after the elections: number of deaths, number of hospitalized patients. All those data were found in governmental data sources, namely the National Institute for Statistics and Economic Studies (INSEE, *Institut National de la Statistique et des Etudes Economiques*) and Public Health France (*Santé Publique France)*. Last, we constructed a specific dataset about participation at the first round of the elections in each French department both in 2014 and 2020. The former date corresponds to the prior version of the same type of election and was deemed necessary not only to measure recent participation but also to compare it in each department with its own predicate. This was made through exhaustive data gathered by IFOP, an international polling institute, city by city, which were then summed to obtain participation rate at the department level.

### Observational analysis

The departments were categorized depending on a composite criterion based on an evaluation of the intensity of the outbreak just before the election, that is from January 24, 2020 through March 18, 2020. This latter date was chosen since it was the closest date for which public data were exhaustively available and since it was assumed that data at this time would also be highly reflective of what was perceived by people at the time of the elections. Three subgroups were created: so-called high intensity departments (more than 70 proven cases and/or more than 25 admitted patients), medium intensity departments (30 to 70 proven cases and/or 10-25 admitted patients) and low intensity departments (less than 30 proven cases and less than 10 admitted patients).

### Statistical analysis and modeling

A sigmoidal mixed effects model was used to describe the cumulative number of admissions for 100 000 habitants: f = *Nmax*\*(exp(*k*\**t*)−1))/(exp(*k*\**t*) + exp*(k* τ*)- Such model has three parameters: the final level *Nmax*, the rate of increase *k*, and the midpoint of increase *τ*. Random effects on the three parameters of the model made it possible to take into account the variability of the number of admissions between departments. Log-normal distributions were used for the three parameters. A covariate analysis then made it possible to analyze how part of the interdepartmental variability of the model parameters could be explained by the participation rates in the 2014 and 2020 elections. Monolix 2019R2 was used to implement and fit the model to the data.

## Results

During the pre-election period, only 278 deaths had been reported in France making this criterion irrelevant for a proper classification of the departments. Generation of the 3 subgroups of departments was based upon the 2954 admitted patients and 4114 cases that were recorded in the pre-election period (Table 1). The more intense was the outbreak, the more populated and denser were the departments (Table 1).

**Table 1.**
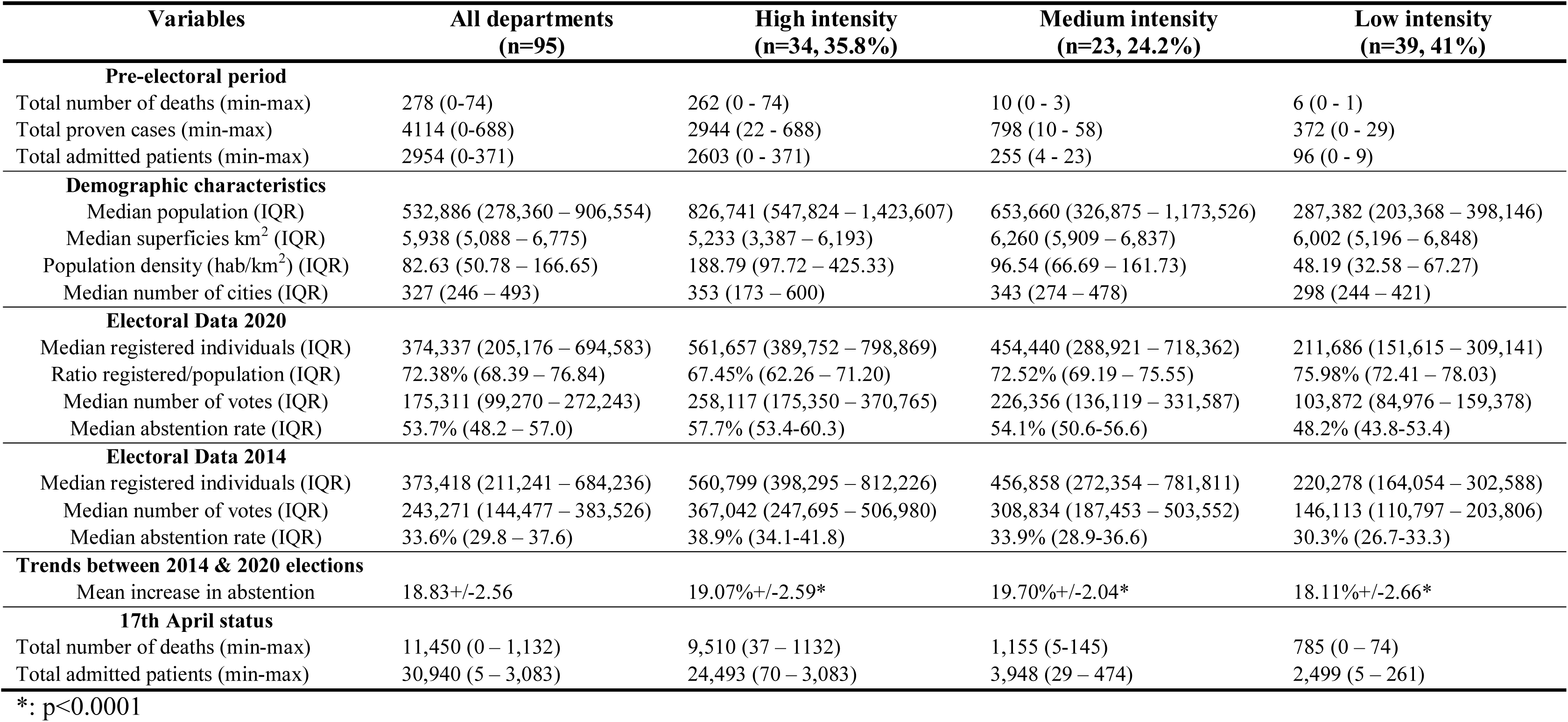
COVID-19 epidemic, demographic data in the pre-electoral period and participation to the vote.

| Variables | All departments<br>(n=95) | High intensity<br>(n=34, 35.8%) | Medium intensity<br>(n=23, 24.2%) | Low intensity<br>(n=39, 41%) |
| --- | --- | --- | --- | --- |
| **Pre-electoral period** |  |  |  |  |
| Total number of deaths (min-max) | 278 (0-74) | 262 (0 - 74) | 10 (0 - 3) | 6 (0 - 1) |
| Total proven cases (min-max) | 4114 (0-688) | 2944 (22 - 688) | 798 (10 - 58) | 372 (0 - 29) |
| Total admitted patients (min-max) | 2954 (0-371) | 2603 (0 - 371) | 255 (4 - 23) | 96 (0 - 9) |
| **Demographic characteristics** |  |  |  |  |
| Median population (IQR) | 532,886 (278,360 – 906,554) | 826,741 (547,824 – 1,423,607) | 653,660 (326,875 – 1,173,526) | 287,382 (203,368 – 398,146) |
| Median superficies km<sup>2</sup> (IQR) | 5,938 (5,088 – 6,775) | 5,233 (3,387 – 6,193) | 6,260 (5,909 – 6,837) | 6,002 (5,196 – 6,848) |
| Population density (hab/km<sup>2</sup>) (IQR) | 82.63 (50.78 – 166.65) | 188.79 (97.72 – 425.33) | 96.54 (66.69 – 161.73) | 48.19 (32.58 – 67.27) |
| Median number of cities (IQR) | 327 (246 – 493) | 353 (173 – 600) | 343 (274 – 478) | 298 (244 – 421) |
| **Electoral Data 2020** |  |  |  |  |
| Median registered individuals (IQR) | 374,337 (205,176 – 694,583) | 561,657 (389,752 – 798,869) | 454,440 (288,921 – 718,362) | 211,686 (151,615 – 309,141) |
| Ratio registered/population (IQR) | 72.38% (68.39 – 76.84) | 67.45% (62.26 – 71.20) | 72.52% (69.19 – 75.55) | 75.98% (72.41 – 78.03) |
| Median number of votes (IQR) | 175,311 (99,270 – 272,243) | 258,117 (175,350 – 370,765) | 226,356 (136,119 – 331,587) | 103,872 (84,976 – 159,378) |
| Median abstention rate (IQR) | 53.7% (48.2 – 57.0) | 57.7% (53.4-60.3) | 54.1% (50.6-56.6) | 48.2% (43.8-53.4) |
| **Electoral Data 2014** |  |  |  |  |
| Median registered individuals (IQR) | 373,418 (211,241 – 684,236) | 560,799 (398,295 – 812,226) | 456,858 (272,354 – 781,811) | 220,278 (164,054 – 302,588) |
| Median number of votes (IQR) | 243,271 (144,477 – 383,526) | 367,042 (247,695 – 506,980) | 308,834 (187,453 – 503,552) | 146,113 (110,797 – 203,806) |
| Median abstention rate (IQR) | 33.6% (29.8 – 37.6) | 38.9% (34.1-41.8) | 33.9% (28.9-36.6) | 30.3% (26.7-33.3) |
| **Trends between 2014 & 2020 elections** |  |  |  |  |
| Mean increase in abstention | 18.83+/-2.56 | 19.07%+/-2.59* | 19.70%+/-2.04* | 18.11%+/-2.66* |
| **17th April status** |  |  |  |  |
| Total number of deaths (min-max) | 11,450 (0 – 1,132) | 9,510 (37 – 1132) | 1,155 (5-145) | 785 (0 – 74) |
| Total admitted patients (min-max) | 30,940 (5 – 3,083) | 24,493 (70 – 3,083) | 3,948 (29 – 474) | 2,499 (5 – 261) |

\*: p<0.0001

Abstention rate in 2020 had a normal distribution among the 95 French departments with a mean of 52.5%±6.4%. Abstention increased by a mean of 18.8% as compared with the 2014 election. There was a significant difference in the variation of abstention between the 3 subgroups of departments (Table 1). Figure 1 represents the abstention rate in the French departments after the 2014 and 2020 elections, as well as the localization of the departments among the three groups. There was a high degree of similarity between 2014 and 2020 with respect to the patterns of departments as compared to others, i.e. regardless of the absolute level of voting, “higher participants” were approximately the same departments at both elections and “lower participants” also. Also, we found a linear correlation among the departments between the abstention rate in 2014 and 2020 (p<0.001).

**Figure 1.**
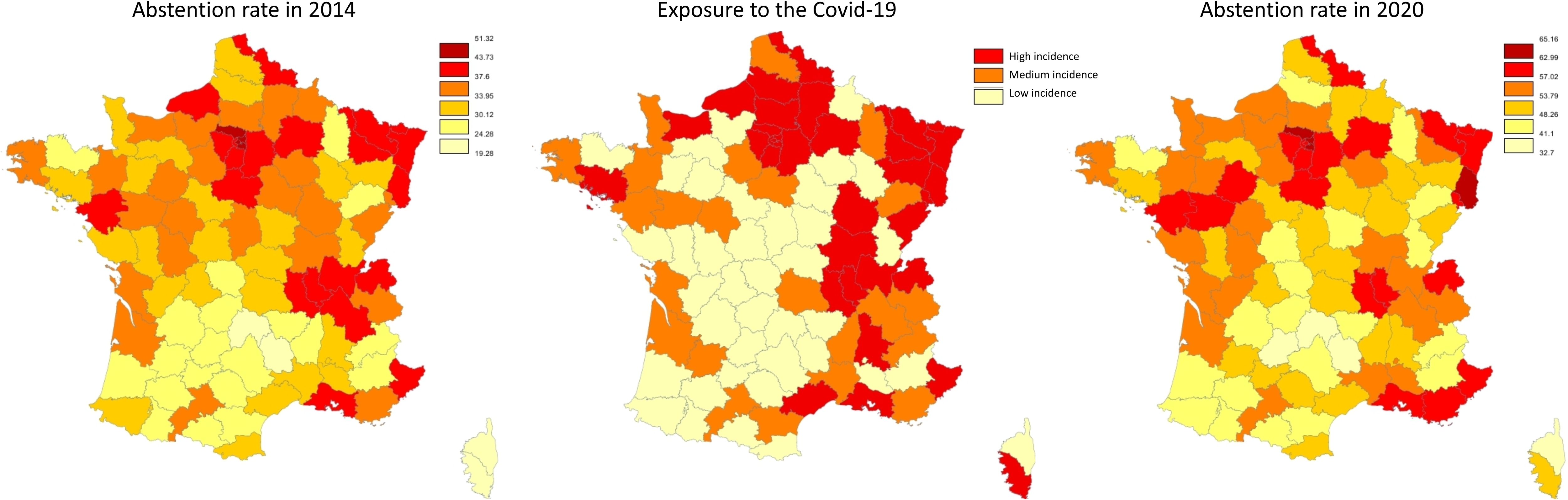
Map of abstention rate among the 95 departments in France after 2014 and 2020 municipal elections and localization of departments among the 3 groups in Covid-19 incidence before 2020 election.

The observed curves of admissions following the election in each subgroup of departments are presented in the Figure 2, with the upper half of departments in terms of participation in red lines and the lower half in blue. Among the departments with a high outbreak intensity, those with a higher participation were not affected by a significantly higher number of admissions for COVID-19 after the elections. Indeed, the departments with a lower participation (i.e. a high abstention) were the ones with more admitted cases. The same divergence was observed in the low intensity departments yet with a smaller difference.

**Figure 2.**
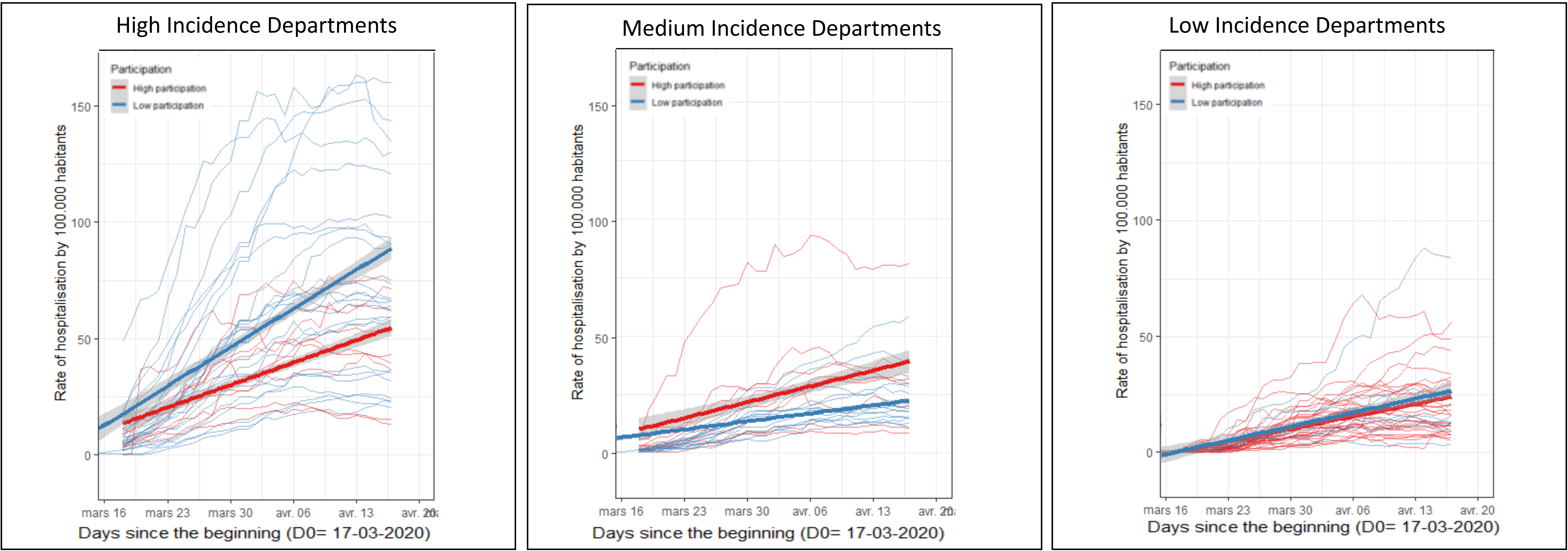
Observed curves of admissions following the election in each subgroup of departments.

### Modeling

Figure 3 showed that the sigmoidal model fitted the data from the different departments with a high degree of consistency. The covariate analysis showed that the 2020 participation explained a very significant part of the variability in the final level *Nmax* (β=-5.36, p<1e^−9^). However, a similar level of significance was obtained with the 2014 participation (β=-3.15, p<1e^−6^), suggesting that the association was most likely due to a confounding variable and not a direct causation specific of the 2020 election.

**Figure 3.**
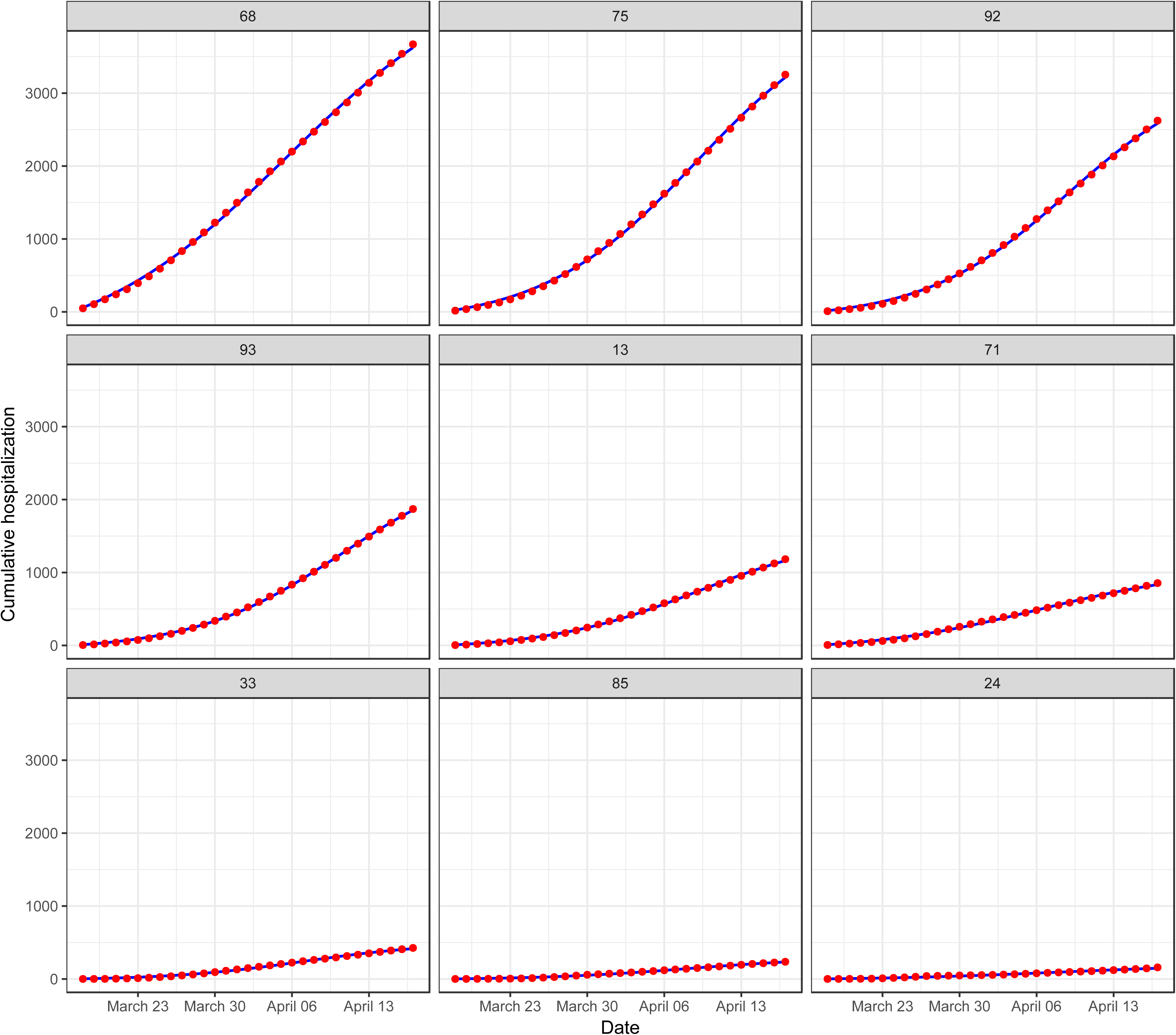
Observed versus predicted cumulative hospitalization number in **9** French department. The red points indicate the observed value, the blue line is the model prediction. The upper panel represent 3 departments among the high incidence group, the medium panel represent 3 departments among the medium incidence group and the lower panel represent 3 departments among the low incidence group.

The difference of participation between the two elections (2020 - 2014) in the covariate model increased slightly the hospitalization rate, but this observed effect was not statistically significant (β=0.096, p=0.35) (Figure 4).

**Figure 4.**
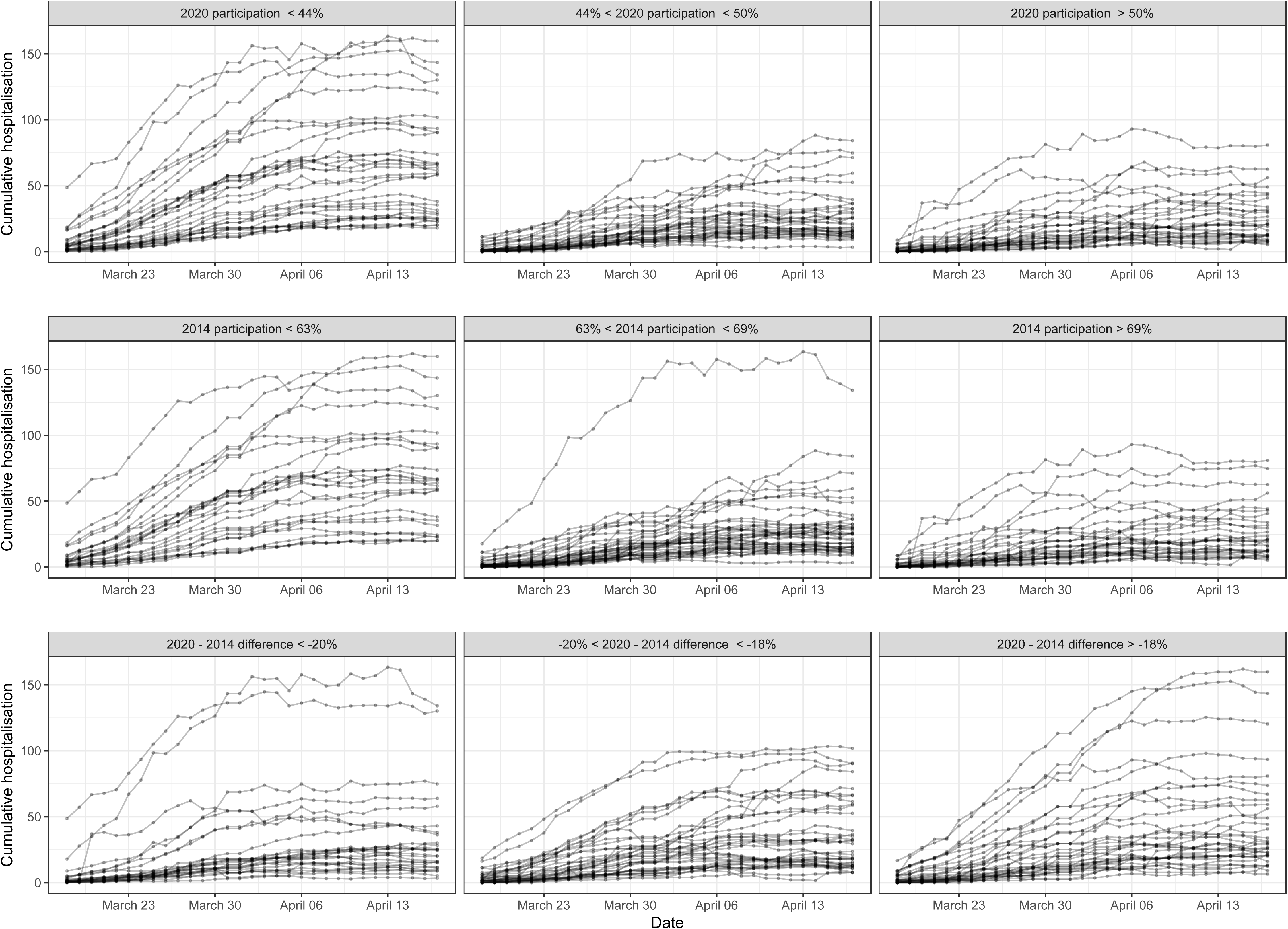
Cumulative hospitalizations over time in tertile of 2020 participation (upper panel), 2014 participation (middle panel) and 2020-2014 differences (lower panel). Each line is a French department with the point representing the observed value.

The relationships between the participation rates and the other two parameters of the model *k* and *τ* were not statistically significant, that participation had no effect on the speed of spread.

## Discussion

Our study is the first national evaluation to date of the reciprocal association between the intensity of the COVID-19 epidemic in France and the level of voting to municipal elections. We did not find any solid evidence suggesting that the level of intensity of the epidemic had a local impact on voting participation. Neither we found any positive statistical association between the level of participation and the subsequent spread of the epidemic in terms of hospitalizations.

Our study has several strengths. First, it is a nationwide analysis gathering exhaustive data regarding all studied variables and outcomes. Patient outcome data, although aggregated at the department level, are thought to be trustworthy since reporting of hospitalizations, as well as in-hospital deaths from COVID-19 were mandatory. Those data were made publicly available and updated on a daily basis by *Santé Publique France*, a public agency depending upon the French Ministry of Health. Data regarding participation to the vote were all collected and structured in an exhaustive manner by IFOP, a polling institute of reference. Then, we chose to retain hospital admissions as an outcome of the possible impact of people participation since it is a clinically relevant endpoint, and since it would allow an earlier assessment as compared to deaths. Also, we used strong and sophisticated methods, such as maximum likelihood estimation, covariate model building and model validation for nonlinear mixed effects models.

Our findings may have implications. The fact that people participation was initially found to be commensurate to the local intensity of the epidemic could have suggested that when prevalent and aggressive, the outbreak deterred citizens to go voting. Importantly, the level of abstention at the national scale was historically low, and more than 30,000 mayors were re-elected after this first round, out of approximately 35,000 cities in the whole territory. [4] However, subsequent accounting for 2014 data showed that there was actually no significant association between the differences in participation between both elections and the local intensity of the outbreak. This in fact suggests that the outbreak may have had a national effect on participation yet independently of local intensity. We also found that on the other hand, while there had been great concern that the holding of the elections might trigger an acceleration of the epidemic spread, there was no positive association between the level of participation and subsequent numbers of admitted patients for COVID-19. Even though it has been reported in the lay press that some so-called assessors (city officials or citizen volunteers participating to the counting of the ballots and to the organization in general) were affected by COVID-19 soon after the holding of the elections, we could not identify any significant effect of the level of participation on following local evolution.

Our report has a main limitation, namely its ecological design. We could not access individual data. We cannot exclude the possibility that our results might be confounded by factors that were not measured and cannot establish (or exclude) definitive causality.

In conclusion, we did not find any local effect of the intensity of the COVID-19 epidemic in France on participation to a national election for mayor designation. While there had been a concern that participation could accelerate virus transmission, we did not measure any statistical association between participation at a local level and subsequent evolution of the epidemic. Our results do not support the election as an interfering factor for outbreak outcome.

## Data Availability

JDZ has access to all the data

## Acknowledgments

None.

## Author contribution

Dr Zeitoun had full access to all of the data in the study and takes responsibility for the integrity of the data and the accuracy of the data analysis.

*Concept, design and acquisition:* Zeitoun, Fourquet, Manternach.

*Statistical analysis:* Lavielle, Faron and Lefevre.

*Analysis and interpretation of data:* Zeitoun, Lavielle, Faron and Lefevre.

*Drafting the manuscript:* Zeitoun.

*Critical revision of the manuscript for important intellectual content:* Zeitoun, Faron and Lefevre.

*Administrative, technical, or material support:* Zeitoun, Fourquet, Mantemach.

